# Large-Language Models for data extraction from written kidney biopsy reports

**DOI:** 10.64898/2026.02.23.26346945

**Authors:** L. Niggemeier, D. L. Hölscher, T. C. J. Herkens, P. Gilles, P. Boor, R. D. Bülow

## Abstract

**Introduction:** Kidney biopsy reports contain rich information that is clinically actionable and useful for research. However, the narrative format hinders scalable reuse. We here investigated whether open-source large language models (LLMs) can extract relevant, standardized readouts from native kidney biopsy pathology reports.

**Methods:** German free-text native kidney biopsy reports were parsed with three open-source LLMs (Llama3 70B, Llama3 8B, MedGemma) to generate structured JSON outputs covering relevant report elements (e.g., diagnosis, glomerular counts, histopathological patterns). Two independent observers manually curated the same report elements; disagreements between the two were resolved by an experienced nephropathologist to create the final ground truth. Performance was assessed using strict and soft matching and summarized accuracy. Inter-rated agreement was quantified using Cohen’s and Light’s Kappa with 95% confidence intervals via 1000-times bootstrapping.

**Results:** Llama3 70B achieved the highest overall accuracy (93.3% strict, 97.1% soft), followed by MedGemma. These larger models showed near perfect performance for explicit and discrete variables and positivity of immunohistochemistry markers, while accuracy decreased for report elements requiring interpretation (e.g., primary diagnosis, interstitial inflammation in fibrosis vs. non-fibrotic cortex). Human raters showed strong agreement for the primary diagnosis (κ = 0.74, 95% CI 0.64-0.84). Adding Llama3 70B or MedGemma as a third rater increased overall agreement (0.82, 95% CI 0.74-0.89 and 0.78, 95% CI 0.69-0.85, respectively), whereas Llama3 8B reduced it.

**Conclusions:** Open-source LLMs can accurately transform narrative nephropathology reports into a structured and machine-readable format, potentially supporting scalable retrospective cohort building. While some report elements can be extracted without supervision, interpretation-dependent elements should be supervised by a human observer.

**Lay Summary:** Retrospective data collection from nephropathology reports is essential for building informative cohorts in computational nephropathology research, yet manual processing of narrative reports is time-consuming and limits scalability. In this study, we demonstrate that open-source large language models can reliably extract key diagnostic, quantitative, and descriptive data elements from kidney biopsy reports with high accuracy. While factual and clearly stated report elements can be extracted automatically, findings that require contextual or interpretative judgment still benefit from expert supervision. Overall, this approach substantially reduces manual effort and enables efficient generation of structured datasets from diagnostic routine, facilitating the development of kidney registries and future computational nephropathology research. In addition, such systems could be implemented into the routine diagnostic workflow, to directly transform narrative reports into structured data.

## Introduction

Kidney biopsies are a cornerstone diagnostic procedure in nephrology, providing essential histopathological information for disease classification, prognosis, and treatment planning. Reporting in nephropathology is mostly free-text-based, which limits comprehensive data extraction and use in research projects. In other fields, particularly cancer pathology, implementation of synoptic and structured reporting has demonstrated clear benefits in clarity, completeness, and data reuse^1,2^. Adoption of these practices holds promise for nephropathology, particularly for standardized kidney biopsy coding and disease registries^3^. Large language models (LLMs) offer new opportunities across many sectors, notably in healthcare^4,5^. By applying advanced natural language processing techniques, LLMs can interpret and analyze complex medical terminology and unstructured data^6^. This includes accurately extracting detailed information from written texts, thereby transforming free-text into structured and machine-readable formats^7^. Previous studies have investigated the potential of LLMs for structured information extraction from radiology and pathology reports^7–9^. These studies did not include the challenging field of nephropathology, characterized by a very large number of rare diseases, nuanced and manifold histological changes, and many specialized stains and imaging techniques such as electron microscopy. One study used natural language processing in nephropathology, but only for the extraction of the main diagnoses^10^.

Here, we show the potential of open-source LLMs for standardized and comprehensive extraction of readouts from kidney biopsy pathology reports. By enabling scalable, precise, and interoperable biopsy data extraction, LLM-driven structured reporting promises to improve research data quality for digital pathology approaches. Because some report elements are inherently interpretative, we additionally quantify inter-rater variability and assess model performance relative to each observer and to consensus labels.

## Short Methods

Narrative native kidney biopsy reports were parsed using three open-source LLMs to extract structured, machine-readable data. The model output was compared to manual curation of the same data performed by two independent observers, with nephropathologists’ adjudication for the final ground truth. Performance was evaluated using strict and soft matching, and summarized via accuracy and inter-rater agreement (Cohen’s/Light’s κ with 95% CIs from 1,000 bootstrap resamples). Full methodological details are available from the supplementary material.

## Results

### Automated report generation

Structured diagnostic data were generated from German free-text histopathological native kidney biopsy reports using three open-source LLMs (Llama3 70B, Llama3 8B, and MedGemma; Figure 1). All models were tasked to extract the same essential relevant elements from written nephropathology reports, including the primary diagnosis, number of glomeruli, number of globally sclerotic glomeruli, relevant histopathological scores and patterns, and immunohistochemical markers (Supp. Table 1), i.e., elements relevant to potential synoptic reports for nephropathology^11^. Each model output was formatted into a standardized pathology report in JSON format for comparison with human-generated references and to support downstream use in kidney databases or computational analysis workflows.

**Figure 1.**
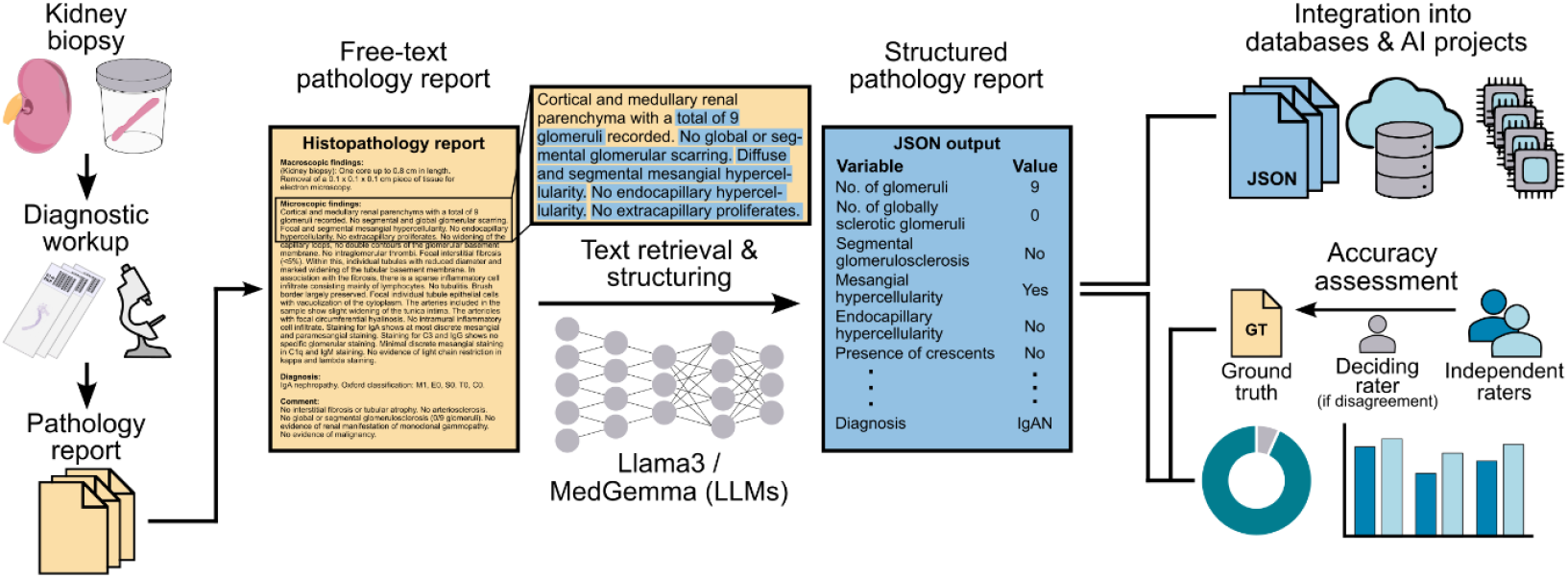
Overview of the data extraction framework using large-language models (LLMs). Unstructured free-text pathology reports from diagnostic routine in a nephropathology center serve as input for LLM-based text retrieval and structuring. The resulting output is a structured JSON report that can be integrated into large databases and AI-driven projects. Model accuracy was assessed by comparing model-generated reports to a human ground truth.

### Model Performance

Based on predefined consensus thresholds, extracted items were categorized as strict, soft or no matches. Strict matches required exact agreement with the ground truth, while soft matches allowed minor deviations, such as incomplete or rephrased expressions. Cases not meeting either criterion were labeled as no match. Additional examples and detailed scoring rules are provided in the Supplementary Methods. Across all evaluated diagnostic categories, Llama3 70B achieved the highest overall accuracy among the tested models (Figure 2a, Supp. Table 1), with an overall accuracy of 93.3% for strict matches and 97.1% for soft matches. MedGemma also performed robustly, with an overall accuracy of 90.5% (strict matches) and 95.9% (soft matches). In contrast, Llama3 8B showed lower accuracy, achieving 79.3% (strict matches) and 84.2% (soft matches).

**Figure 2.**
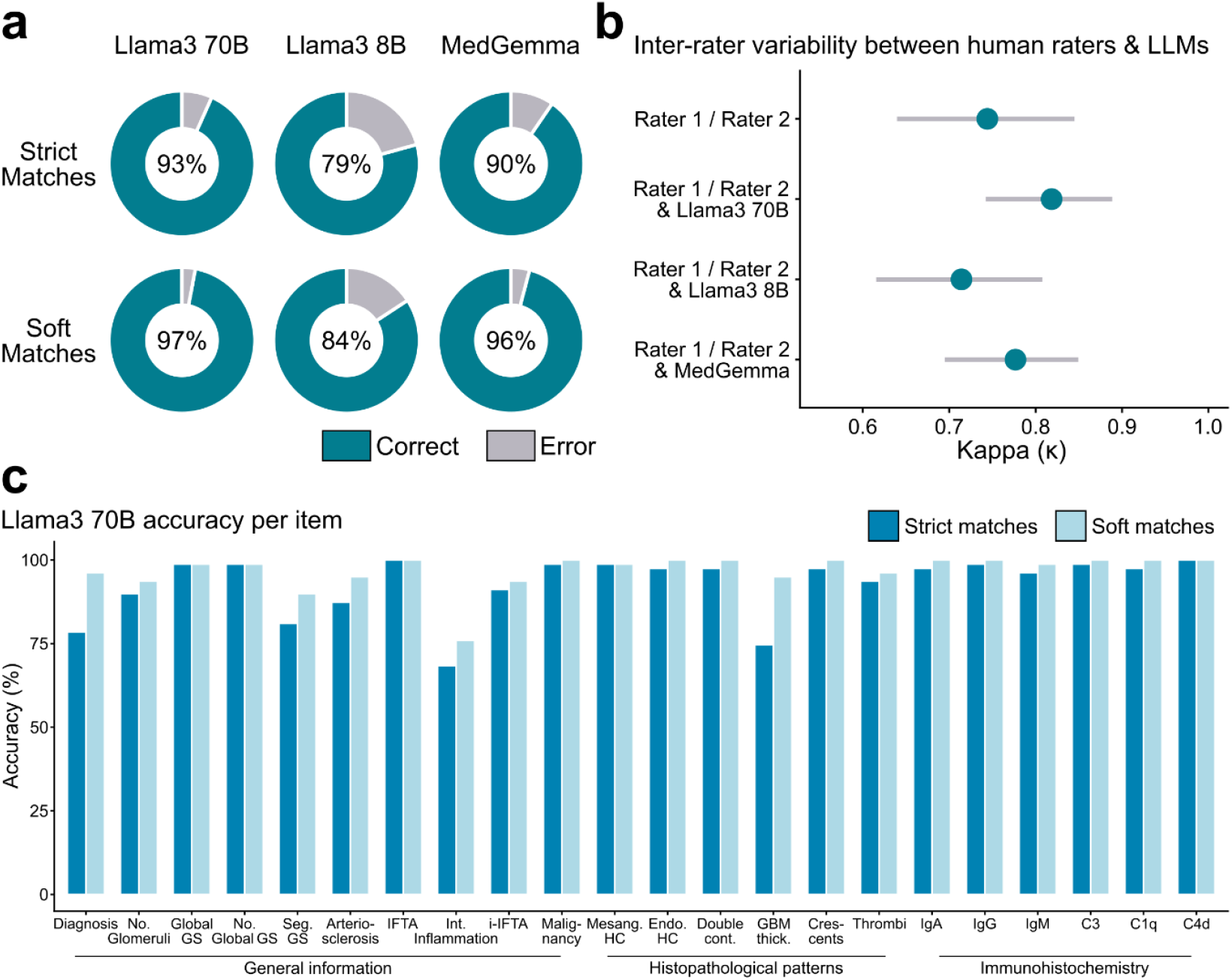
Comparative performance of large-language models (LLM) for data extraction from nephropathology reports. **(a)** Overall accuracy of the three LLMs based on strict and soft matches with the human ground truth. **(b)** Inter-rater variability between two human raters and between both the human raters and each LLMs. **(c)** Item-wise accuracy of Llama3 70B across all sections of a nephropathology report. Abbreviations: No., number, GS, glomerulosclerosis; Segment., segmental; IFTA, interstitial fibrosis and tubular atrophy; Mesang., mesangial; HC, hypercellularity; Endocap., endocapillary; cont, contours; GBM, glomerular basement membrane; thick., thickening; IgA, immunoglobulin A; IgG, immunoglobulin G; IgM, immunoglobulin M.

For individual diagnostic items, Llama3 70B and MedGemma consistently reached near-perfect accuracy (above 95%) across histopathological patterns and immunohistochemistry markers (Figure 2c, Supp. Table 1). Lower accuracy was seen for more context-dependent findings, including the overall diagnosis, interstitial inflammation, and interstitial inflammation in interstitial fibrosis and tubular atrophy (i-IFTA), particularly in the smaller Llama3 8B model (Supp. Table 1). Specialized prompts for specific items partially mitigated these errors. A prompt emphasizing the distinction between interstitial inflammation and i-IFTA increased interstitial inflammation accuracy to 88.6% (+12.6% for soft matches) while i-IFTA accuracy remained high (Supplementary Table 2). In contrast, diagnosis-specific prompts only resulted in an overall increase in accuracy for strict matches (+7.6%), but not for soft matches. A detailed breakdown of representative errors across different items is provided in Table 1.

**Table 1.** Representative examples of errors and ambiguities observed in responses from large language models and human raters.

| Item | Free-text report | Ground truth | Model output | Vote |
| --- | --- | --- | --- | --- |
| Number of glomeruli | No global or segmental glomerulosclerosis (0/8). | 8 | <u>Llama3 70B</u>: Not mentioned | False |
|  |  |  | <u>Llama3 8B</u>: Not mentioned | False |
|  |  |  | <u>MedGemma</u>: Not mentioned | False |
| Diagnosis | Focal segmental glomerulosclerosis with focal segmental glomerular foam cells and global podocyte injury. | Focal segmental glomerulosclerosis (1 human rater voted “Not mentioned” as the diagnosis was understood as a pattern) | <u>Llama3 70B</u>: Focal segmental glomerulosclerosis | Strict match |
|  |  |  | <u>Llama3 8B</u>: Focal segmental glomerulosclerosis with focal segmental glomerular foam cells and global podocyte injury. | Soft match |
|  |  |  | <u>MedGemma</u>: Focal segmental glomerulosclerosis | Strict match |
| Diagnosis | Focal segmental glomerulosclerosis with focal global glomerulosclerosis (1 of 7 glomeruli) and global podocyte injury. | Focal segmental glomerulosclerosis | <u>Llama3 70B</u>: Focal segmental glomerulosclerosis | Strict match |
|  |  |  | <u>Llama3 8B</u>: Focal segmental glomerulosclerosis and focal global glomerulosclerosis | Soft match |
|  |  |  | <u>MedGemma</u>: Focal segmental glomerulosclerosis | Strict match |
| Diagnosis | Diffuse global glomerulosclerosis (8 of 12 glomeruli). Ultrastructurally, electron-dense deposits with a fibrillar substructure. Taking into account the electron microscopic finding of electron-dense deposits with a fibrillar substructure (12–20 nm in diameter) that stain positively for immunoglobulins (IgG dominant) and complement, the findings are consistent with a fibrillary glomerulonephritis. | Fibrillary Glomerulonephritis | <u>Llama3 70B</u>: Fibrillary Glomerulonephritis | Strict match |
|  |  |  | <u>Llama3 8B</u>: Diffuse global glomerulosclerosis (8 of 12 glomeruli). Ultrastructurally, electron-dense deposits with a fibrillar substructure. | False |
|  |  |  | <u>MedGemma</u>: Diffuse global glomerulosclerosis | False |
| Arterio-sclerosis | The arteries show moderate to focally severe thickening of the tunica intima. Up to severe arteriosclerosis. | Severe arteriosclerosis | <u>Llama3 70B</u>: Moderate arteriosclerosis | False |
|  |  |  | <u>Llama3 8B</u>: Moderate arteriosclerosis | False |
|  |  |  | <u>MedGemma</u>: Moderate<br>arteriosclerosis | False |

### Human and LLM inter-rater variability

Because nephropathology reports often contain nuanced information, we assessed inter-rater variability between the two human raters and after adding each LLM as a third rater for the primary diagnosis. We observed strong agreement between both independent human raters (Cohen’s κ = 0.74, 95% CI 0.64-0.84; Figure 2b). Including the LLM-specified diagnosis as a third rater improved overall agreement for Llama3 70B (Light’s κ = 0.82, 95% CI 0.74-0.89) and MedGemma (Light’s κ = 0.78, 95% CI 0.69-0.85), but it decreased for Llama3 8B (Light’s κ = 0.71, 95% CI 0.62-0.81; Figure 2b). Notably, agreement between human raters and between human raters and LLMs tended to decrease for diagnostically ambiguous terminology, such as cases where focal segmental glomerulosclerosis (FSGS) could be interpreted either as a descriptive histopathological pattern or as a glomerular disease (Table 1). Extracting structured data using the best performing model LLama3 70B was 12,5-17,86-times faster than human data collection.

## Discussion

We here evaluated whether open-source LLMs can transform narrative nephropathology reports from native kidney biopsies into structured, machine-readable data suitable for downstream research use. Across a broad set of report elements, the strongest open LLMs achieved high concordance with human-extracted reference labels, showing particularly strong performance for explicitly stated, discrete variables (e.g., glomerular counts and clearly enumerated markers). In contrast, performance decreased for interpretation-dependent elements (e.g., inflammation in relation to fibrosis).

Retrospective collection of such structured nephropathology datasets remains a major barrier to computational nephrology research projects. In many centers, reporting is predominantly free text^11^, and manual data extraction is time-consuming and not scalable. However, scalable approaches are urgently needed to facilitate research, particularly for data-driven AI research requiring large datasets^12^. Our results suggest that LLM-based extraction can substantially accelerate retrospective structuring of reports and may enable more rapid creation of analysis-ready datasets.

A key observation is that some aspects of the report, particularly those requiring integration of multiple phrases or mapping descriptive patterns to diagnostic categories, showed lower agreement and higher observer variability. This issue was partially improved by specialized prompts, although not for all items. Our results support using LLMs as scalable second observers and pre-extraction tools: low-risk, explicitly stated variables can be auto-populated with high confidence, whereas high-risk interpretive variables may be pre-filled but likely require refined prompts or targeted verification.

Future work should include multicenter and multilingual validation, prospective evaluation in routine workflows, and mapping extracted fields to controlled vocabularies, such as the Kidney Biopsy Codes to improve interoperability. In addition, studying the impact of structured extraction on downstream endpoints, such as registry completeness, cohort finding accuracy, and reproducibility of computational analyses, will clarify where LLM-based structuring provides the greatest real-world value in computational nephropathology.

## Data Availability

Code supporting this study is publicly available in the Zenodo repository 14175293 and was adapted from a previous study of Grothey et al.7,13. The model prompts used for all LLMs are provided as Supplementary Material. Due to privacy constraints, kidney biopsy reports cannot be made publicly available; however, access may be granted upon reasonable request to the corresponding author, in such cases, a data transfer agreement approved by the local legal department and ethical approval is required.

## Data and code availability

Code supporting this study is publicly available in the Zenodo repository 14175293 and was adapted from a previous study of Grothey et al.^7,13^. The model prompts used for all LLMs are provided as Supplementary Material. Due to privacy constraints, kidney biopsy reports cannot be made publicly available; however, access may be granted upon reasonable request to the corresponding author, in such cases, a data transfer agreement approved by the local legal department and ethical approval is required.

## Ethics statement

Data collection and analysis in this study was performed in accordance with the Declaration of Helsinki and was approved by the local ethics committee of the RWTH Aachen University (EK-No. 125/25).

## Author contributions

Conceptualization: LN, DLH, PB, RDB; Methodology: LN, DLH, RDB; Software: LN, DLH, RDB; Data Curation: TCJH, PG, RDB; Formal analysis: LN, DLH, RDB; Resources: PB, RDB; Writing, Original Draft: LN; Review & Editing: LN, DLH, TCJH, PG, PB, RDB; Visualization: LN, DLH; Supervision: PB, RDB; Project administration: None; Funding acquisition: PB, RDB.

