## Supplementary Material for "Large-Language Models for data extraction from written kidney biopsy reports"

### Supplementary Methods

#### Report extraction

Written reports were extracted from the local Laboratory Information System (LIS, Nexus Pathology) by querying the database using the following search restrictions: “Materialart”: “Eigenniere”, translating to “Specimen type”: “Native Kidney” with the inclusion period 01.11.2022 to 31.12.2025. Written reports were downloaded in batches due to software restrictions and the written content of the reports were extracted and transferred to a csv file using custom PowerShell code. Reports from light microscopy as well as reports from electron microscopy were included. From a randomly selected time period, we included 80 consecutive nephropathology reports for human scoring and model assessment. One report was excluded due to inconsistencies, resulting in a total of 79 nephropathology reports.

#### Large-language model tuning

Three open-source models were selected for information extraction and automated report generation: a large and small Llama3 model with 70 billion and 8 billion parameters[1], respectively, and a midsize model, MedGemma, with 27 billion parameters[2]. Both Llama3 models were trained on general text data while MedGemma was specifically trained on medical data. All models were inferred using the *pipeline* function of the *Huggingface transformers* library[3]. The implementation approach was adapted from a previous study of Grothey et al.[4,5] (Zenodo repository: 14175293). The same model prompt was used for all LLMs (provided as supplementary material). All models were run under deterministic decoding ensuring reproducibility. Specifically, all parameters governing sampling-based randomness were disabled or set to limiting values (e.g., temperature = 0, no top-k sampling), ensuring deterministic next-token prediction. Inference was conducted on four NVIDIA DGX A100 80GB GPUs.

#### Performance evaluation

Model performance was assessed using a ground truth established by two independent raters, i.e., medical students with previous experience in nephropathology. In case of disagreement between both raters, a third independent rater, i.e., a trained nephropathologist, was consulted for a final decision. The third rater was blinded to the model output when reaching his decision. For all three models, each extracted information was checked for correctness, with two agreement thresholds. Strict matches required exact agreement between model output and ground truth, with the exception of small formatting changes, e.g., *"less than 5%"* vs. *"<5%"* or *"2.0"* vs. *"2"*. Soft matches permitted minor deviations such as rephrased or incomplete expressions, e.g., *"Focal segmental"* vs. *"Yes"* for presence of mesangial hypercellularity or *"low-grade"* vs. *"minimal"* for arteriosclerosis.

#### Prompt improvement strategies

To improve performance of two items with more nuanced reporting and lower model accuracy, we tested specialized prompts for each item. The first item was diagnosis. Here, the only task for the LLM is to extract the relevant diagnosis from the opinion section of the report. Since we observed many issues regarding reporting FSGS as a diagnosis even though only the pattern was described in the report, we gave the hint that FSGS is not a diagnosis, except if it appears in the context of a podocytopathy. Additionally, the LLM was instructed to only extract diseases as diagnosis but not histological patterns. The second category was interstitial inflammation. Here, we observed mix-ups with i-IFTA. In the specialized prompt, we gave the task to only extract information about interstitial inflammation and i-IFTA with the information to properly distinguish between the two. The full prompts are provided as Supplementary Material.

#### Statistical analysis

All analyses and statistical calculations were conducted in R and Python. Model accuracy was calculated as the percentage of correctly classified items and summarized across all evaluated categories to obtain an overall model accuracy. Inter-rater variability for the primary diagnosis of each case was assessed using Cohen's Kappa (κ) for pairwise human comparisons and Light's Kappa (κ) for multi-rater analyses including both human and model raters. 95% confidence intervals for Kappa values were derived through nonparametric bootstrapping with 1,000 resamples.

### Supplementary Tables

**Supplementary Table 1.** Overall item-wise accuracy for strict and soft matches across all three large-language models.

| | **Model** | **Llama3 70B** | | **Llama3 8B** | | **MedGemma** | |
|---|---|---|---|---|---|---|---|
| **Item** | **Threshold** | Strict | Soft | Strict | Soft | Strict | Soft |
| Diagnosis | | 78.5% | 96.2% | 69.6% | 82.3% | 58.2% | 91.1% |
| No. of glomeruli | | 89.9% | 93.7% | 54.4% | 54.4% | 84.8% | 87.3% |
| Global glomerulosclerosis | | 98.7% | 98.7% | 94.9% | 94.9% | 98.7% | 100.0% |
| No. of globally sclerotic glomeruli | | 98.7% | 98.7% | 54.4% | 70.9% | 91.1% | 94.9% |
| Segmental glomerulosclerosis | | 81.0% | 89.9% | 70.9% | 70.9% | 82.3% | 91.1% |
| Arteriosclerosis | | 87.3% | 94.9% | 77.2% | 93.7% | 83.5% | 96.2% |
| IFTA | | 100.0% | 100.0% | 65.8% | 100.0% | 94.9% | 100.0% |
| Interstitial inflammation | | 68.4% | 76.0% | 67.1% | 67.1% | 68.4% | 77.2% |
| i-IFTA | | 91.1% | 93.7% | 78.5% | 78.5% | 93.7% | 96.2% |
| Malignancy | | 98.7% | 100.0% | 92.4% | 100.0% | 100.0% | 100.0% |
| Mesangial hypercellularity | | 98.7% | 98.7% | 91.1% | 92.4% | 98.7% | 100.0% |
| Endocapillary hypercellularity | | 97.5% | 100.0% | 89.9% | 91.1% | 98.7% | 100.0% |
| Double contours | | 97.5% | 100.0% | 88.6% | 88.6% | 97.5% | 100.0% |
| GBM thickening | | 74.7% | 94.9% | 67.1% | 68.4% | 64.6% | 97.5% |
| Crescents | | 97.5% | 100.0% | 83.5% | 83.5% | 96.2% | 100.0% |
| Thrombi | | 93.7% | 96.2% | 89.9% | 89.9% | 96.2% | 96.2% |
| IgA | | 97.5% | 100.0% | 88.6% | 89.9% | 94.9% | 96.2% |

| | | | | | | |
|---|---|---|---|---|---|---|
| IgG | 98.7% | 100.0% | 93.7% | 96.2% | 93.7% | 97.5% |
| IgM | 96.2% | 98.7% | 86.1% | 94.9% | 96.2% | 97.5% |
| C3 | 98.7% | 100.0% | 93.7% | 98.7% | 96.2% | 97.5% |
| C1q | 97.5% | 100.0% | 86.1% | 93.7% | 96.2% | 98.7% |
| C4d | 100.0% | 100.0% | 86.1% | 87.3% | 100.0% | 100.0% |
| No. of biopsy cores | 98.7% | 98.7% | 67.1% | 67.1% | 92.4% | 92.4% |
| Max. length of biopsy core | 100.0% | 100.0% | 79.3% | 84.2% | 93.7% | 93.7% |

**Supplementary Table 2**. Improvements using specialized prompts for three items.

| **Item** | Diagnosis | | Interstitial Inflammation | | i-IFTA | |
|---|---|---|---|---|---|---|
| **Threshold** | Strict | Soft | Strict | Soft | Strict | Soft |
| Original prompt | 78.5% | 96.2% | 68.4% | 76.0% | 91.1% | 93.7% |
| Specialized prompt | 86.1% | 96.2% | 83.5% | 88.6% | 92.4% | 92.4% |
| Difference | +7.6% | +0.0% | +15.1% | +12.6% | +1.3% | -1.3% |

### General Prompt

Please extract key information from the provided pathology report and categorize it under the respective headings. Listed below are the parameters that should be retrieved from the report. All of them are formatted as follows:

Heading: [Possible options]

Stick precisely to the given answer options for each category. If an answer option specifies 'Integer', ensure the response is provided as an integer value. If an answer option specifies 'Float', ensure the response is provided as a float value. If the report does not specify information about a particular category or if the information is unclear, provide the response as "Not mentioned".

The output should be structured in a JSON format.

1. Diagnosis: [short diagnosis/Not mentioned]

2. Number of total glomeruli: [Integer/Not mentioned]

3. Number of globally sclerotic glomeruli: [Integer/Not mentioned]

4. Segmental glomerulosclerosis: [Yes/No/Not mentioned]

5. Malignancy: [Yes/No/Not mentioned]

6. Degree of arteriosclerosis: [weak/moderate/strong/Not mentioned]

7. Interstitial fibrosis and tubular atrophy (IFTA): [Range/Percentage/Not mentioned]

8. Globale glomerulosclerosis: [Yes/No/Not mentioned]

9. Mesangial hypercellularity: [Yes/No/Not mentioned]

10. Endocapillary Hypercellularity: [Yes/No/Not mentioned]

11. Double contours: [Yes/No/Not mentioned]

12. Thickening of the glomerular basement membrane: [Yes/No/Not mentioned]

13. Extracapillary proliferates or crescents: [Yes/No/Not mentioned]

14. Thrombi: [Yes/No/Not mentioned]

15. Interstitial inflammation: [Yes/No/Not mentioned]

16. Interstitial inflammation in fibrosis (iIFTA): [Yes/No/Not mentioned]

17. IgG immunohistochemistry positive: [Yes/No/Not mentioned]

18. IgA immunohistochemistry positive: [Yes/No/Not mentioned]

19. IgM immunohistochemistry positive: [Yes/No/Not mentioned]

20. C3 immunohistochemistry positive: [Yes/No/Not mentioned]

21. C1q immunohistochemistry positive: [Yes/No/Not mentioned]

22. C4d immunohistochemistry positive: [Yes/No/Not mentioned]

23. Number of biopsy cores: [Integer/Not mentioned]

24. Maximum length of biopsy cores in cm: [Float/Not mentioned]

25. Electron microscopic or ultrastructural findings present: [Yes/No/Not mentioned]

26. Electron microscopic or ultrastructural findings requested: [Yes/No/Not mentioned]

27. Further reports regarding this case following: [Yes/No/Not mentioned]

The report reads:

*Followed by the report.*

### Specialized Prompt (1)

#### Diagnosis

Please extract key information from the provided pathology report and categorize it under the respective headings. Listed below are the parameters that should be retrieved from the report. All of them are formatted as follows:

Heading: [Possible options]

Stick precisely to the given answer options for each category. If the report does not specify information about a particular category or if the information is unclear, provide the response as "Not mentioned".
If possible extract the relevant diagnosis in a short format from the opinion section of the report. Focal segmental glomerulosclerosis is not a diagnosis except it is primary with podocytopathy. Only diseases should be used for diagnosis. Histological patterns should not count. This is really important.
The output should be structured in a JSON format.

1. Diagnosis: [diagnosis/Not mentioned]

The report reads:

*Followed by the report.*

### Specialized Prompt (2)

#### Interstitial Inflammation & i-IFTA

Please extract key information from the provided pathology report and categorize it under the respective headings. Listed below are the parameters that should be retrieved from the report. All of them are formatted as follows:

Heading: [Possible options]

Stick precisely to the given answer options for each category. If the report does not specify information about a particular category or if the information is unclear, provide the response as "Not mentioned".
Make sure to properly distinguish between interstitial inflammation and interstitial inflammation in fibrosis. This is an important difference.
The output should be structured in a JSON format.

1. Interstitial inflammation: [Yes/No/Not mentioned]

2. Interstitial inflammation in fibrosis: [Yes/No/Not mentioned]

The report reads:

*Followed by the report.*
